# Deep Learning Prediction of Parkinson’s Disease using Remotely Collected Structured Mouse Trace Data

**DOI:** 10.1101/2024.10.27.24316195

**Authors:** Md Rahat Shahriar Zawad, Zerin Nasrin Tumpa, Lydia Sollis, Shubham Parab, Peter Washington

## Abstract

Parkinson’s Disease (PD) is the second most common neurodegenerative disorder globally, and current screening methods often rely on subjective evaluations. We developed deep learning-based classification models using structured mouse trace data collected via a web application. 261 participants (73 PD, 155 non-PD, 33 suspected PD) completed three hand movement tasks: tracing a straight line, spiral, and sinewave. We developed three types of models: (1) engineered features model, (2) computer vision models, and (3) multimodal models. The best-performing models were image-based DenseNet-201 model with an F1 score of 0.9027 ± 0.0332 (PD vs. non-PD), multimodal ResNet-50 with an F1 score of 0.9353 ± 0.0334 (suspected PD vs. non-PD), and multimodal ViT with an F1 score of 0.7619 ± 0.0535 (PD vs non-PD). Feature importance for the best-performing models was evaluated using Gradient Shapley Additive Explanations (GradShap). Image inputs consistently proved most predictive. The findings suggested that models trained on confirmed PD diagnoses hold promise for early-stage PD screening.

## Introduction

Parkinson’s Disease (PD) is a neurodegenerative disorder that significantly impacts the central nervous system. Major symptoms include tremors, bradykinesia, muscle rigidity, and postural instability^1,2^, which progressively worsen over time, leading to difficulties in performing routine tasks such as typing and using a mouse. The progression of these symptoms significantly affects quality of life, making early and accurate diagnosis crucial to enable early intervention^3^. PD is the second most common neurodegenerative disease after Alzheimer’s, affecting approximately 10 million people globally. In the United States, around one million individuals are diagnosed with PD, with an annual increase of about 90,000 new cases. This number is projected to rise to 1.2 million by 2030^3,4^. Currently, there is no definitive biomarker for PD, and diagnosis is primarily based on clinical symptoms and neuropsychological tests such as the Mini-Mental State Examination (MMSE) and the Unified PD Rating Scale (UPDRS)^5,6,7^. These tests involve questionnaires and subjective evaluations by clinicians, which can lead to significant biases and potential misdiagnoses^6^. This is particularly problematic, as PD symptoms often overlap with those of other age-related conditions and Drug-Induced Parkinsonism (DIP)^8,9,10^. Additionally, PD is primarily caused by the degeneration of dopamine-producing neurons in the brain, and by the time motor symptoms become apparent, approximately 30-50% of these neurons have already deteriorated^11^. Therefore, early and accurate detection of PD is essential for effective management and treatment.

Previous digital health research on PD classification included the analysis of hand and finger movements, keystroke dynamics, speech, handwriting, drawing tests, and sensor data from accelerometers and gyroscopes^12–25^. The use of sensors placed on lower limbs, wearable sensors supported by video recordings, and sensing coils or paper-based pads have proven effective in classifying PD and detecting tremors^16–28^. However, these methods often require controlled laboratory settings and specialized devices, limiting their broader applicability. Self-administered methods, such as keyboard interactions, keystroke dynamics, and smartphone screen interactions, have also been explored but may introduce biases, particularly against individuals with slower typing speeds^13,14,20^. Mobile applications for data collection, symptom monitoring, and treatment management have demonstrated utility in tracking activities like finger tapping speed, gait, and motor performance, though they pose challenges for older adults unfamiliar with smartphone technology^29,30^. Multimodal approaches combining data from speech, gait, and upper limb movements have demonstrated potential in classifying PD patients but often require controlled environments and specialized equipment, which limits their use in real-world contexts^31,32^.

We aim to address these challenges and advance the field of digital PD screening by utilizing structured mouse trace data collected through a short test delivered on a user-friendly web application. Participants recruited for the study provided demographic information and completed tasks involving the tracing of straight line, spiral, and sine wave patterns using their mouse. We performed feature engineering on the collected mouse trace data and created images from the mouse movement patterns to develop computer vision models. We trained a simple feedforward neural network for analyzing engineered features. For the mouse trace images, we utilized state-of-the-art computer vision models including Vision Transformer (ViT), Shifted Window Transformer (SwinT), DenseNet-201, ResNet-50, and MobileNet V2. To enhance classification performance, we developed multimodal models that combined the engineered features with the mouse trace images. We analyzed the performance of the models on three different sets of train-test splits: the first set included PD and non-PD in both train and test data, the second set included PD and non-PD for training while suspected PD and non-PD for testing, and the third set used suspected PD and non-PD for training while PD and non-PD for testing.

## Methods

We created a website to collect structured mouse movement data by having participants complete a series of mouse tracing tests remotely. We trained a series of deep learning models using various features derived from the mouse traces. **Figure 1** outlines the workflow from data collection to model evaluation and interpretability analysis.

**Figure 1.**
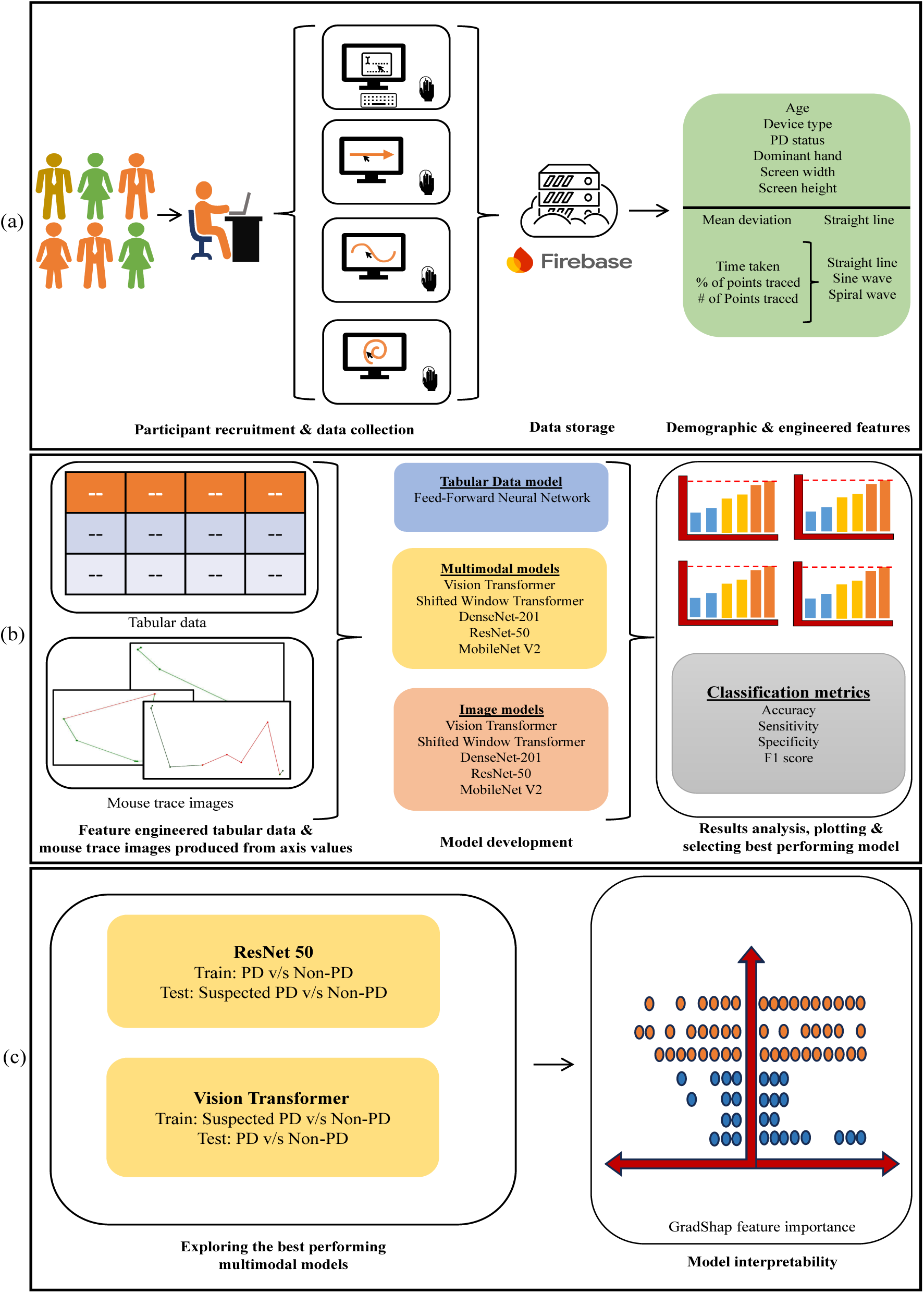
The study workflow from data collection to model evaluation and interpretability analysis. (a) Participants completed the data collection process remotely on the website. (b) Engineered features and mouse trace images were fed into three sets of models. The results were analyzed and visualized to identify the best performing models. (c) The best multimodal models (multimodal ResNet-50 and multimodal ViT) were analyzed for interpretability using GradShap feature importance.

### Ethics approval

The study was approved by the University of Hawaii Institutional Review Board (IRB, protocol #2023-00948).

### Participant Recruitment & Data Collection

We recruited participants for this study through both online methods (email, social media posts) and offline methods (community meetings and conventions in Hawaii). We collaborated with the Hawaii Parkinson Association and Beyond Rehab to post flyers and advertise the study to various PD e-mailing lists. Additionally, we established a recruitment booth at the 2024 Hawaii Parkinson’s Association Symposium and 2024 Hawaii Annual Parkinson’s Run, where we provided potential participants with flyers describing how to complete the study.

We collected data via a web application that we developed (https://parkinsonsurvey.github.io/), illustrated in **Figure 2**. Participants provided demographic and disease-related information, including age, and dominant hand. Due to the absence of official diagnostic documents and biomarkers for PD, self-reporting was used, with an option to select "suspected PD".

**Figure 2:**
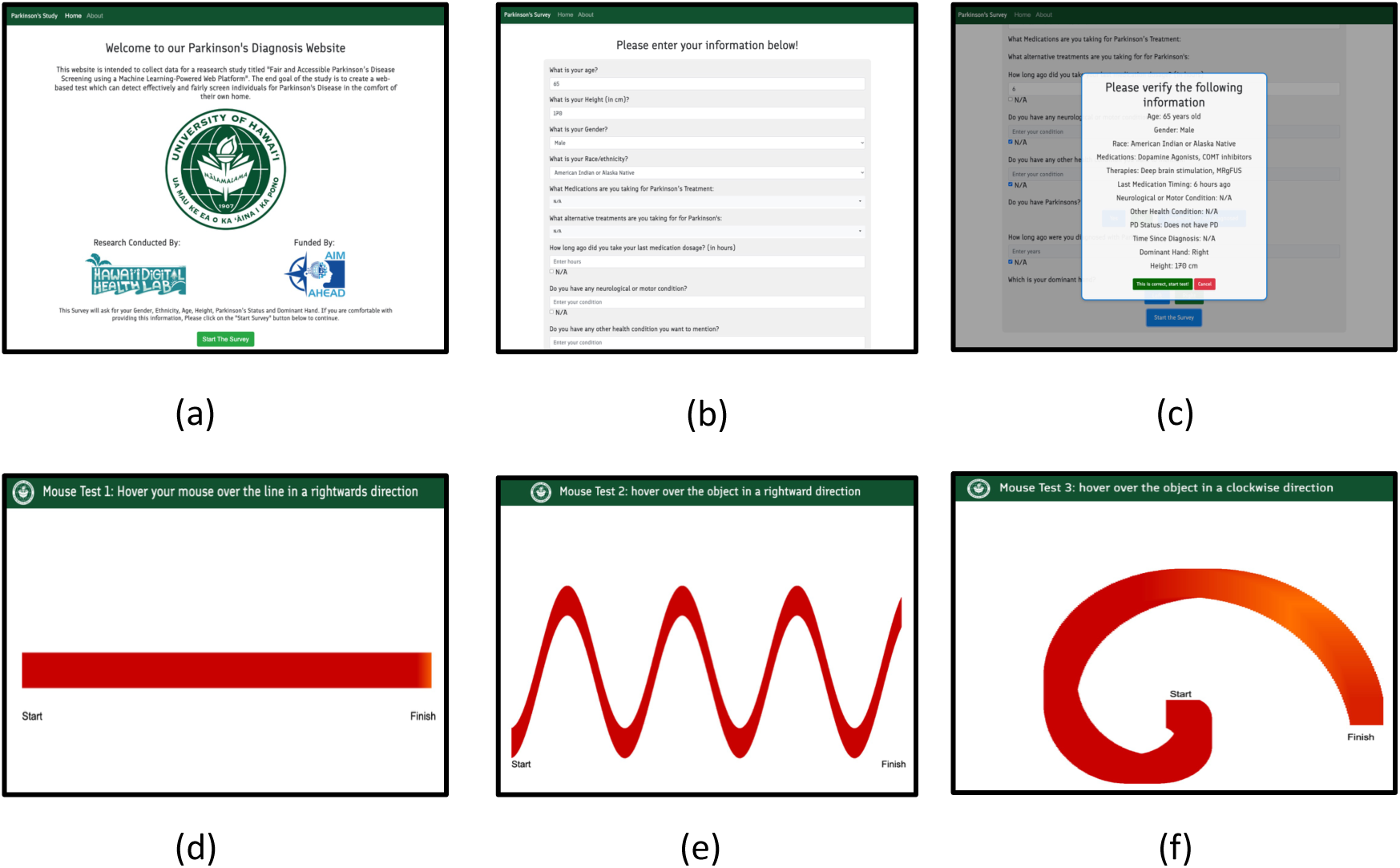
Pages of the data collection website: (a) Introduction page of the website to inform the participants about the study. (b) The participants were asked to provide information about themselves. (c) Participants were asked to confirm their information to prevent mistakes. (d) Participants were asked to trace a straight line. (e) Participants were asked to trace a sine wave. (f) Participants were asked to trace a spiral wave.

Participants used a physical mouse on their desktop or laptop, to trace a straight line, sine wave, and spiral wave on the website. We visualized their progress and alignment with the lines through highlighted portions and start/end markings. We developed the website using HTML and Bootstrap for the interface and visuals, and JavaScript to track cursor position every 500 milliseconds. The data included mouse position (X, Y axis), and whether the mouse was inside the line (True or False). The web application also captured screen dimensions and operating system details for contextual information. Upon completing the test, all data were securely transmitted and stored in a Firebase collection.

### Feature Engineering & Mouse Trace Image Generation

Based on the raw data collected, we engineered several features to quantify tracing performance across three tracing tasks (straight-line, sine wave, and spiral). These features include:

- *Mean Deviation from the Line (Straight-line only):* The average absolute vertical deviation of the traced points from the expected centerline, normalized by screen height. The centerline is set based on the screen height and adjusted to match the web interface.
- *Time Taken* (milliseconds): The total tracing time is approximated by multiplying the number of recorded points by the 500 milliseconds sampling interval.
- *Time Taken with Respect to Window Width* (milliseconds per pixel): The total time normalized by the screen width, calculated as the total time divided by the window width. This metric accounts for variations in screen size between participants.
- Percentage of Points Traced Inside the Target: The proportion (from 0 to 1) of recorded points that fall inside the expected trace area.
- *Raw Count of Points Traced Inside the Target:* Simply the number of points recorded inside the expected trace area.

Using these features shown in **Table 1**, we quantified the consistency and accuracy of a participant’s trace on each test pattern.

**Table 1.** Engineered features extracted from mouse trace data.

| Feature Name | Description | Applicable Trace |
| --- | --- | --- |
| **Mean Deviation from Centerline (fraction of screen height)** | Average absolute vertical deviation from the expected centerline, normalized by the screen height. | Straight-line |
| **Time Taken (milliseconds)** | Total time estimated by multiplying the number of recorded points by 500 ms. | All |
| **Time Taken with Respect to Window Width (ms per pixel)** | Total time divided by the window width. This normalizes the tracing duration to account for differences in screen dimensions. | All |
| **Percentage of Points Traced Inside (as a fraction, 0 to 1)** | Percentage of the number of points inside the expected trace region to the total number of points. | All |
| **Number of Points Traced Inside** | Count of points that are within the expected trace region, irrespective of timing. | All |

To generate mouse trace images, we created canvases corresponding to each participant’s screen dimensions. The recorded x and y coordinates were then used to render the trace, where segments starting with points inside the target region are colored green, and those that fall outside are colored red. **Figure 3** illustrates samples of these plots.

**Figure 3:**
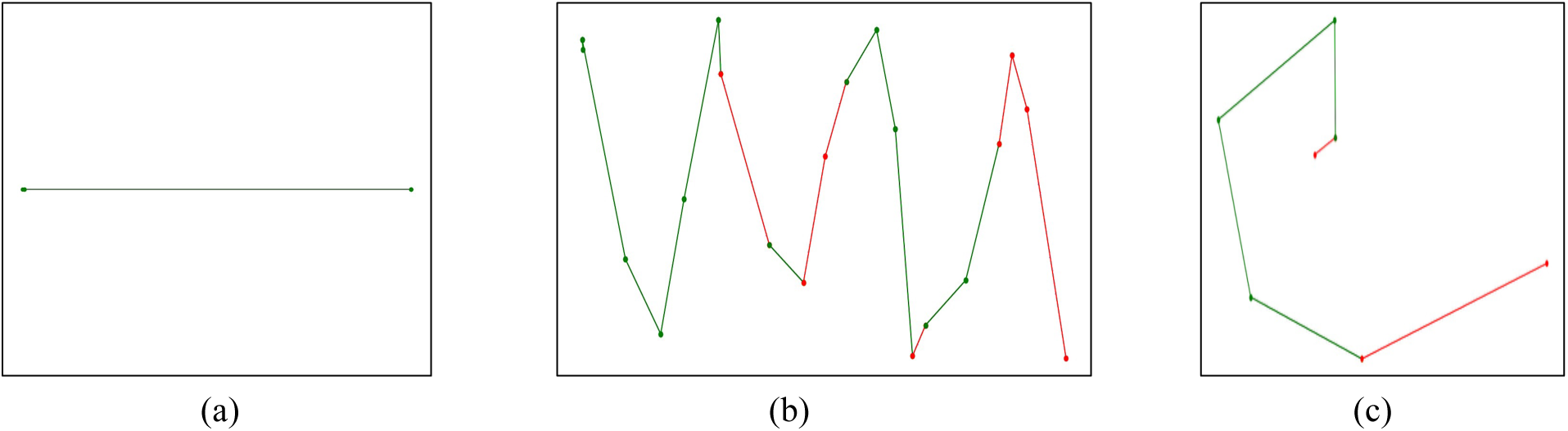
Samples of mouse trace images produced: (a) Stright line. (b) Sine wave. (c) Spiral Wave.

**Figure 4.**
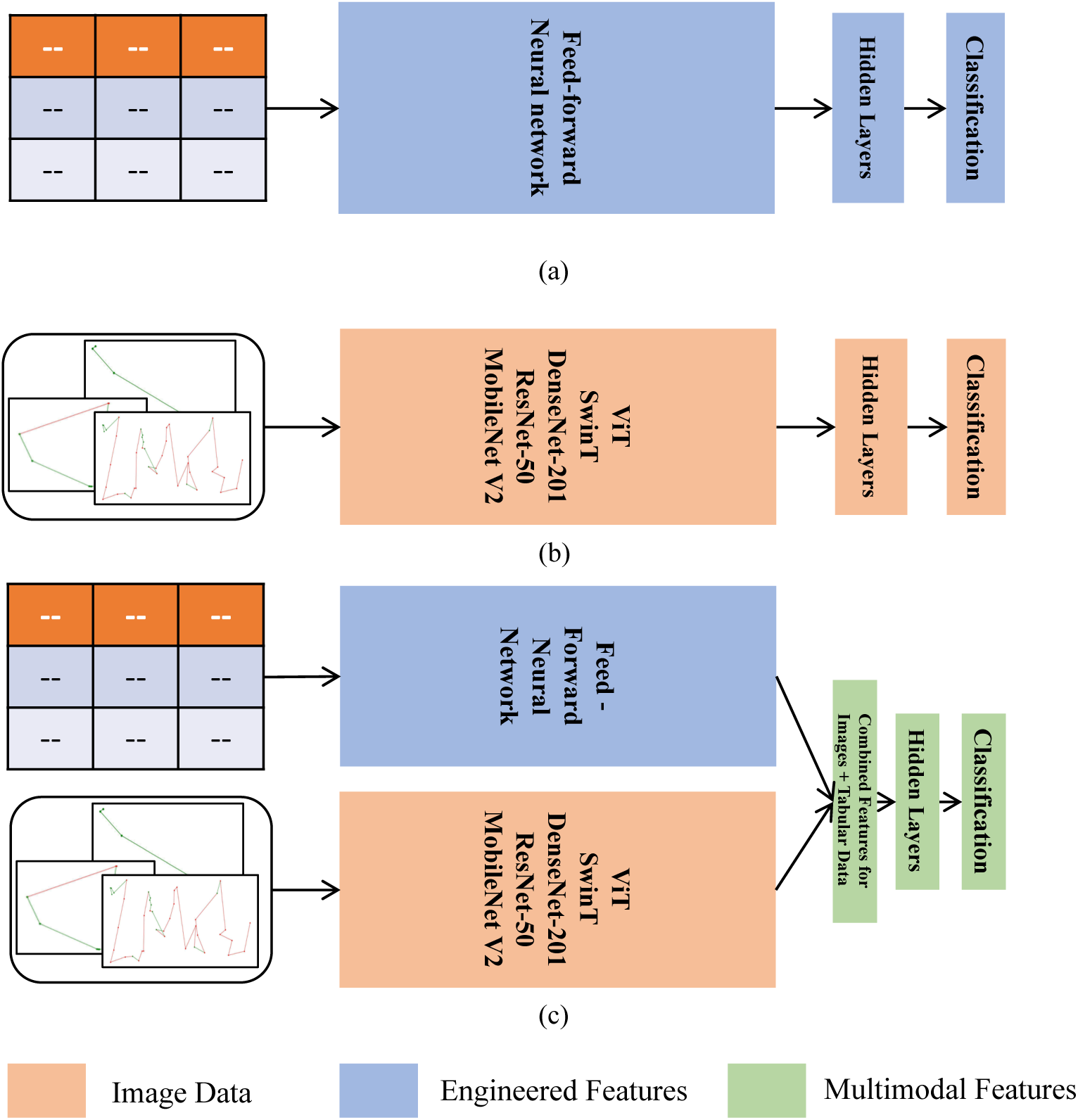
Model Architectures. (a) Tabular data models. (b) Image data models. (c) Multimodal models.

### Model Development

We developed three categories of deep learning models (**Figure 3.4**), each utilizing different input modalities, to better understand the individual and combined contributions of image and tabular data to PD detection. The first category focused on engineered tabular features, processed through a feed-forward neural network. The second category comprised image-based models, where input images were passed through pre-trained architectures including ViT, SwinT, DenseNet-201, ResNet-50, and MobileNet V2. Transfer learning was applied by unfreezing the final 45 layers of each network and replacing the classification heads to match our binary labels. The third category consisted of multimodal models, which integrated both data types: tabular features were passed through feed-forward neural network, while image inputs were processed via the same pre-trained backbones used in the image-only models. The resulting feature representations were concatenated and passed through additional hidden layers for classification. All models were trained as binary classifiers for up to 50 epochs, with early stopping applied using a patience of 5. By structuring the models consistently across these categories, we aimed to isolate and analyze the impact of each data modality and understand how individual components contribute to the performance of the multimodal architectures.

### Data Splitting and Hyperparameter Tuning

We evaluated our models using three distinct train-test splits designed to assess different aspects of model generalization:

In split 1, we included only participants labeled as either PD or non-PD, explicitly excluding those marked as suspected PD. Models were trained and evaluated using 5-fold cross-validation. Within each fold, hyperparameters were tuned using Optuna across 10 configurations, and performance was estimated using 500 bootstrapped resamples per fold. To mitigate variability caused by non-deterministic behavior observed in several of our image-based and multimodal models, we repeated each fold 10 times with independently initialized models, reporting average performance across runs. This experiment established a baseline for distinguishing confirmed PD from controls.

In split 2, the training set comprised all confirmed PD participants and 60% of the non-PD samples, while the test set and validation set included all suspected PD participants combined with the remaining 40% of non-PD data equally divided. This setup aimed to evaluate whether models trained on confirmed PD cases could detect early or ambiguous motor patterns in suspected PD individuals. Hyperparameters were tuned using Optuna over 30 trials, and final performance was estimated by averaging results across 20 independently trained models, each evaluated using 500 bootstrap resamples.

In split 3, models were trained on all suspected PD participants along with 60% of the non-PD data and evaluated on all confirmed PD cases plus the remaining 40% of non-PD samples equally divided in test and validation set. This experiment assessed whether models trained on borderline or early-stage patterns could generalize to detect confirmed PD. As in Experiment 2, 20 independent training runs were conducted to account for non-deterministic effects, and evaluation was performed using 500 bootstrap resamples per run.

However, we reported single run results for the models with deterministic behavior from experiment 2 and 3 so that the variability from bootstrapping can be traced on those models.

### Evaluation Metrics

To evaluate model performance, we used four commonly used classification metrics: sensitivity, specificity, accuracy, and F1 score. These were calculated using the confusion matrix derived from model predictions, consisting of:

- *True Positives (TP)*: cases correctly predicted as positive.
- *True Negatives (TN)*: cases correctly predicted as negative.
- *False Positives (FP)*: negative cases incorrectly predicted as positive.
- *False Negatives (FN)*: positive cases incorrectly predicted as negative.

From these values we calculated, Sensitivity, the proportion of actual positive cases correctly identified, defined as 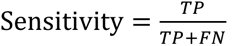. Specificity, the proportion of actual negative cases correctly classified, defined as 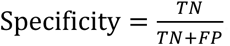. Accuracy, the overall proportion of correctly classified cases across both classes, calculated as 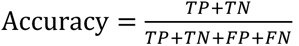. F1 score, the harmonic mean of precision and recall, particularly useful for imbalanced class distributions and is computed as 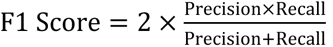. Where, 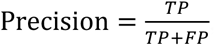, and 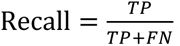.

Each of these metrics was estimated using 500 bootstrapped resamples to compute both the mean and standard deviation, providing a robust measure of model stability and generalizability. We used F1 score as the base metric to select the best performing models from our analysis.

### Saliency Map & Feature Importance

To identify important features for the best-performing model architectures, we created GradShap saliency plots, as GradSHAP is optimized for identifying complex feature importances in deep learning models^33^. GradSHAP was applied to both the image and engineered tabular inputs to quantify their respective contributions to the model’s predictions. For the image modality, attributions were computed separately for the sine, straight, and spiral images by comparing the model’s output on actual inputs versus shuffled versions of the same batch, used as baselines. Attribution values were summed across spatial dimensions to yield a single importance score for each image input. Importantly, the sign of each attribution was preserved to indicate the direction of influence: positive values reflect contributions toward a PD or suspected PD prediction, while negative values indicate support for a non-PD classification.

## Results

### Dataset

We had 261 participants with completed attempts for the mouse trace section of the website. Among the 261 participants, 73 self-reported as having PD, 155 as non-PD, and the remaining 33 as suspected PD. Attempts with data available for all three mouse trace tasks irrespective of the age group were included. The characteristics of the collected data is shown in **Figure 5** and **Table 2**.

**Figure 5.**
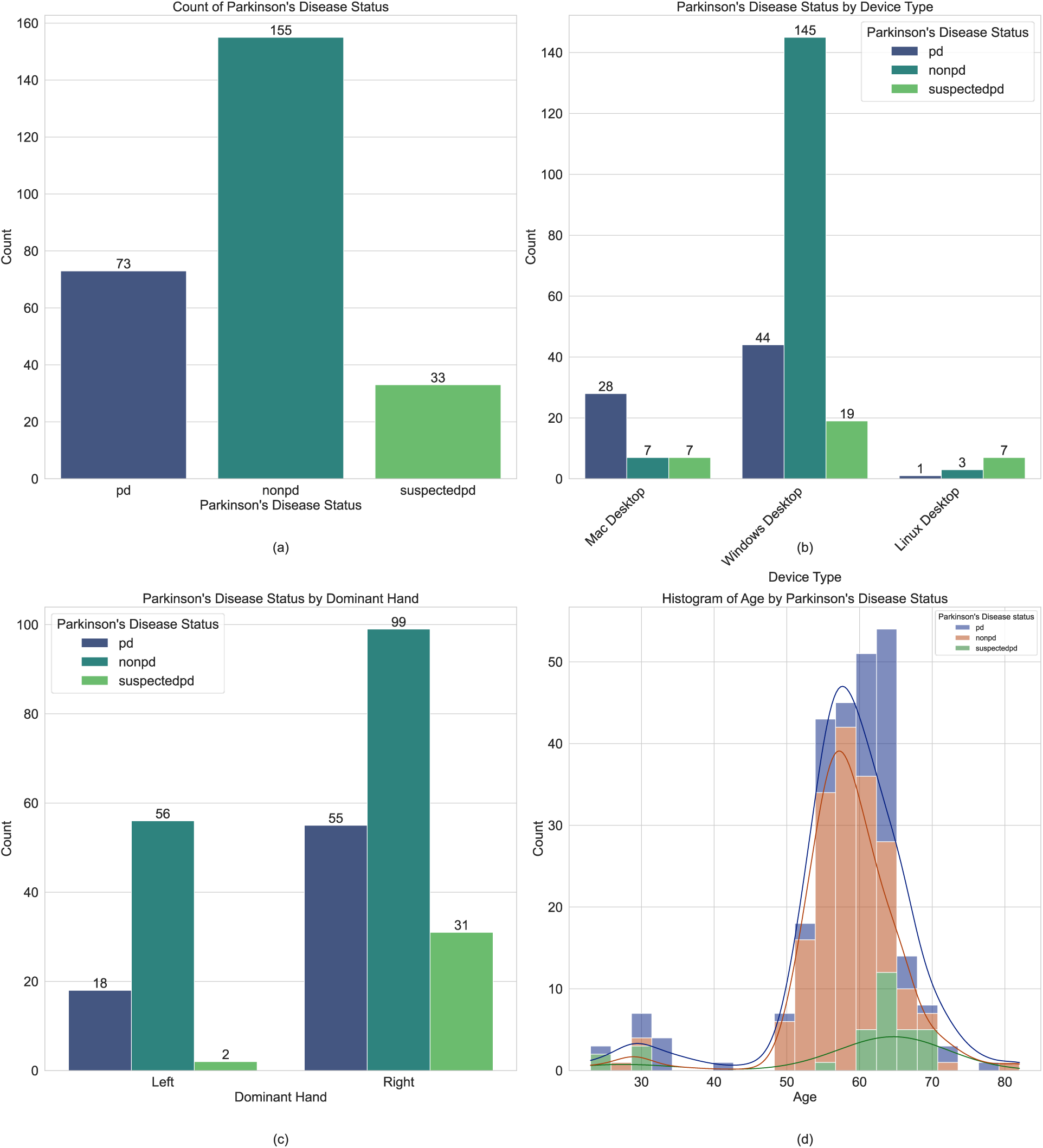
Participant distribution among the different classes: (a) PD, non-PD, and suspected PD, stratified by: (b) device type, (c) dominant hand and, (d) age.

**Table 2.** Participant distribution among the different classes (PD, non-PD, and suspected PD) for age, dominant hand, and device type.

| Characteristics | Participants | PD | Non-PD | Suspected PD |
| --- | --- | --- | --- | --- |
| **Age** |  |  |  |  |
| 23 - 49 | 16 | 9 | 2 | 5 |
| 50 - 59 | 113 | 15 | 97 | 1 |
| 60 - 69 | 126 | 46 | 53 | 27 |
| 70 + | 6 | 3 | 3 | 0 |
| **Dominant Hand** |  |  |  |  |
| Right | 185 | 55 | 99 | 31 |
| Left | 76 | 18 | 56 | 2 |
| **Device type** |  |  |  |  |
| Windows | 208 | 44 | 145 | 19 |
| Mac | 42 | 28 | 7 | 7 |
| Linux | 11 | 1 | 3 | 7 |
| **Total** | 261 | 73 | 155 | 33 |

### Performance Metrics

**Figure 6.**
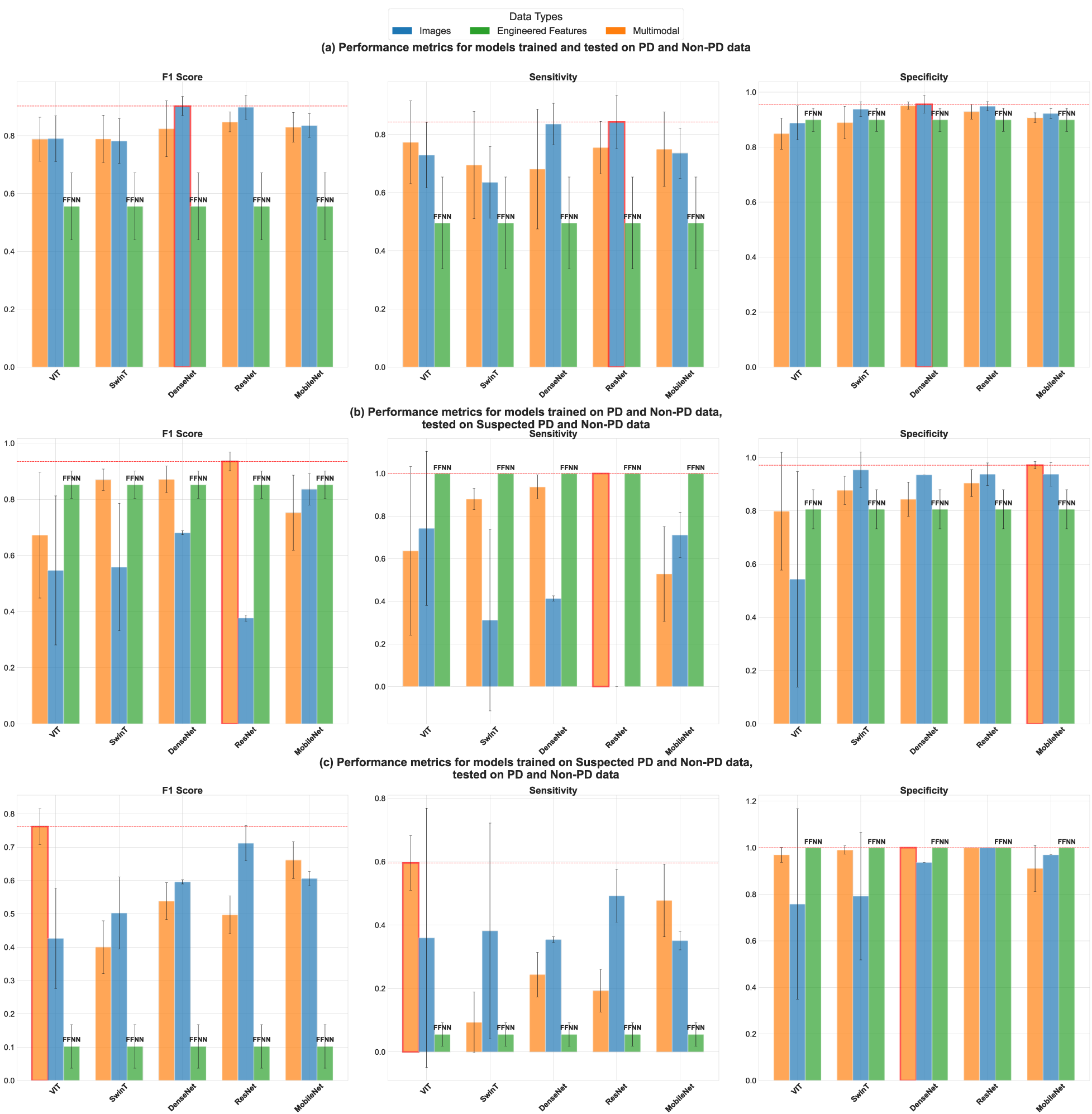
Key performance metrics (F1-score, sensitivity, and specificity) for all models. (a) Trained and Tested on PD vs Non-PD data (on 5 fold cross validation), (b) Trained on PD vs Non-PD, tested on Suspected PD vs Non-PD data and, (c) Trained on Suspected PD vs Non-PD, tested on PD vs Non-PD data evaluated with 500 resampling bootstraps. The error bars represent standard deviation values.

#### 5-Fold Cross Validation Predicting PD vs Non-PD

In our first split, using PD and non-PD data for both training and testing via 5-fold cross validation, the image-based DenseNet-201 model achieved the highest F1 score (0.9027 ± 0.0332), alongside an accuracy of 0.9179 ± 0.0288, sensitivity of 0.8360 ± 0.0713, and specificity of 0.9557 ± 0.0327. Overall, the pre-trained CNNs (ResNet-50: F1 = 0.8986 ± 0.0413; MobileNet V2: F1 = 0.8355 ± 0.0410; and DenseNet-201: F1 = 0.9027 ± 0.0332) outperformed both transformer-based image models (ViT: F1 = 0.7900 ± 0.0789; SwinT: F1 = 0.7819 ± 0.0774) and the engineered-feature only feed-forward neural network (F1 = 0.5558 ± 0.1161), which showed high specificity (0.8993 ± 0.0419) but low sensitivity (0.4955 ± 0.1583). When tabular features were added to the image data, multimodal SwinT saw a very small F1 gain (around +0.0071). In contrast, all other multimodal models experienced F1 drops of approximately 0.001 to 0.0785. These results suggest that while image-based CNNs capture the majority of diagnostic signals from mouse trace images, the benefit of integrating engineered features is almost nonexistent for most of the architectures. Complete performance metrics for this split are provided in **Table 3**, and **Figure 6(a)** plots F1, sensitivity, and specificity for all models.

**Table 3.** The performance metrics for models trained and tested on PD and non-PD data; evaluated using 5-fold cross validation with 500 bootstraps resampling. All values are mean ± standard deviation.

| Model | Data Type | Accuracy | Sensitivity | Specificity | F1 |
| --- | --- | --- | --- | --- | --- |
| ViT | Multimodal | $0.8255 \pm 0.0529$ | $0.7730 \pm 0.1422$ | $0.8488 \pm 0.0566$ | $0.7883 \pm 0.0757$ |
| SwinT | Multimodal | $0.8263 \pm 0.0608$ | $0.6949 \pm 0.1847$ | $0.8891 \pm 0.0595$ | $0.7890 \pm 0.0822$ |
| DenseNet 201 | Multimodal | $0.8654 \pm 0.0638$ | $0.6809 \pm 0.2060$ | $0.9507 \pm 0.0134$ | $0.8242 \pm 0.0968$ |
| ResNet-50 | Multimodal | $0.8727 \pm 0.0260$ | $0.7547 \pm 0.0905$ | $0.9286 \pm 0.0266$ | $0.8476 \pm 0.0341$ |
| MobileNet V2 | Multimodal | $0.8566 \pm 0.0367$ | $0.7492 \pm 0.1275$ | $0.9071 \pm 0.0175$ | $0.8294 \pm 0.0513$ |
| ViT | Images | $0.8366 \pm 0.0545$ | $0.7290 \pm 0.1128$ | $0.8875 \pm 0.0619$ | $0.7900 \pm 0.0789$ |
| SwinT | Images | $0.8404 \pm 0.0421$ | $0.6352 \pm 0.1230$ | $0.9379 \pm 0.0270$ | $0.7819 \pm 0.0774$ |
| DenseNet 201 | Images | $0.9179 \pm 0.0288$ | $0.8360 \pm 0.0713$ | $0.9557 \pm 0.0327$ | $0.9027 \pm 0.0332$ |
| ResNet-50 | Images | $0.9141 \pm 0.0329$ | $0.8430 \pm 0.0917$ | $0.9484 \pm 0.0168$ | $0.8986 \pm 0.0413$ |
| MobileNet V2 | Images | $0.8629 \pm 0.0319$ | $0.7359 \pm 0.0865$ | $0.9220 \pm 0.0183$ | $0.8355 \pm 0.0410$ |
| FFNN | Engineered features | $0.7689 \pm 0.0432$ | $0.4955 \pm 0.1583$ | $0.8993 \pm 0.0419$ | $0.5558 \pm 0.1161$ |

### Training on PD vs Non-PD, testing on Suspected PD vs Non-PD

In our second split, models were trained on PD and non-PD data and tested on suspected PD and non-PD data. The multimodal ResNet-50 model achieved the highest F1 score (0.9353 ± 0.0334), with balanced sensitivity (1.0000 ± 0.0000) and specificity (0.9045 ± 0.0506). The feed-forward neural network using only engineered features scored an F1 of 0.8523 ± 0.0488 (accuracy = 0.8746 ± 0.0474, sensitivity = 1.0000 ± 0.0000, specificity = 0.8058 ± 0.0734), showing that tabular data alone still carries useful signal but may not match multimodal models in this case. Among the image-based models, MobileNet V2 was the best, with an F1 of 0.8359 ± 0.0560 (accuracy = 0.8575 ± 0.0470, sensitivity = 0.7118 ± 0.1058, specificity = 0.9375 ± 0.0443). DenseNet-201 followed with F1 = 0.6809 ± 0.0073 (accuracy = 0.7507 ± 0.0043, sensitivity = 0.4137 ± 0.0122, specificity = 0.9355 ± 0.0000), while SwinT and ViT trailed (SwinT F1 = 0.5588 ± 0.2272; ViT F1 = 0.5467 ± 0.2659). Notably, ResNet-50 trained on images failed to detect any positives (sensitivity = 0.0000 ± 0.0000) and scored only F1 = 0.3769 ± 0.0108. Adding engineered features with image inputs produced positive effects for F1 score: ResNet-50 jumped from 0.3769 ± 0.0108 to 0.9353 ± 0.0334, DenseNet-201 jumped from 0.6809 ± 0.0073 to 0.8712 ± 0.0472, SwinT jumped from 0.5588 ± 0.2272 to 0.8697 ± 0.0380, and ViT went from 0.5467 ± 0.2659 to 0.6726 ± 0.2242. In contrast, MobileNet V2’s F1 score dipped from 0.8359 ± 0.0560 to 0.7523 ± 0.1336. These results indicate that while detecting early or ambiguous motor patterns in suspected PD, multimodal fusion can improve robustness and performance balance. Complete performance metrics for this split are shown in **Table 4**, and comparisons of F1 score, sensitivity, and specificity are visualized in **Figure 6(b)**.

**Table 4.** The performance metrics for models trained on PD and non-PD data, tested on suspected PD and non-PD data with 500 bootstraps resampling. All values are mean ± standard deviation.

| Model | Data Type | Accuracy | Sensitivity | Specificity | F1 |
| --- | --- | --- | --- | --- | --- |
| ViT | Multimodal | $0.7414 \pm 0.1585$ | $0.6370 \pm 0.3958$ | $0.7986 \pm 0.2211$ | $0.6726 \pm 0.2242$ |
| SwinT | Multimodal | $0.8782 \pm 0.0369$ | $0.8801 \pm 0.0501$ | $0.8771 \pm 0.0528$ | $0.8697 \pm 0.0380$ |
| DenseNet 201 | Multimodal | $0.8768 \pm 0.0465$ | $0.9371 \pm 0.0561$ | $0.8438 \pm 0.0645$ | $0.8712 \pm 0.0472$ |
| ResNet-50 | Multimodal | $0.9383 \pm 0.0327$ | $1.0000 \pm 0.0000$ | $0.9045 \pm 0.0506$ | $0.9353 \pm 0.0334$ |
| MobileNet V2 | Multimodal | $0.8145 \pm 0.0747$ | $0.5285 \pm 0.2227$ | $0.9713 \pm 0.0138$ | $0.7523 \pm 0.1336$ |
| ViT | Images | $0.6135 \pm 0.2308$ | $0.7422 \pm 0.3615$ | $0.5430 \pm 0.4047$ | $0.5467 \pm 0.2659$ |
| SwinT | Images | $0.7263 \pm 0.1106$ | $0.3119 \pm 0.4256$ | $0.9535 \pm 0.0675$ | $0.5588 \pm 0.2272$ |
| DenseNet 201 | Images | $0.7507 \pm 0.0043$ | $0.4137 \pm 0.0122$ | $0.9355 \pm 0.0000$ | $0.6809 \pm 0.0073$ |
| ResNet-50 | Images | $0.6054 \pm 0.0274$ | $0.0000 \pm 0.0000$ | $0.9374 \pm 0.0424$ | $0.3769 \pm 0.0108$ |
| MobileNet V2 | Images | $0.8575 \pm 0.0470$ | $0.7118 \pm 0.1058$ | $0.9375 \pm 0.0443$ | $0.8359 \pm 0.0560$ |
| FFNN | Engineered features | $0.8746 \pm 0.0474$ | $1.0000 \pm 0.0000$ | $0.8058 \pm 0.0734$ | $0.8523 \pm 0.0488$ |

### Training on Suspected PD vs Non-PD, testing on PD vs Non-PD

In our third split, where models trained on suspected PD and non-PD data were tested on confirmed PD and non-PD cases, the multimodal ViT model performed best, achieving an F1 of 0.7619 ± 0.0535 with balanced performance (sensitivity = 0.5961 ± 0.0859, specificity = 0.9694 ± 0.0319 and accuracy = 0.7663 ± 0.0500). Overall, performance rose as we moved from tabular to image data: the feed-forward neural network (F1 = 0.1022 ± 0.0654) achieved only 0.0551 ± 0.0369 in sensitivity but 1.000 ± 0.000 in specificity. Looking at image-only models, ResNet-50 led the group (F1 = 0.7119 ± 0.0529; accuracy = 0.7236 ± 0.0455; sensitivity = 0.4921 ± 0.0836; specificity = 1.0000 ± 0.0000), while DenseNet-201 and MobileNet V2 performed lower (DenseNet-201 F1 = 0.5958 ± 0.0062; MobileNet V2 F1 = 0.6059 ± 0.0216). Adding engineered features to the image backbones produced mixed results for F1 score: ViT’s F1 score jumped from 0.4263 ± 0.1504 to 0.7619 ± 0.0535, MobileNet V2 went from 0.6059 ± 0.0216 to 0.6611 ± 0.0549, but F1 score for DenseNet201, SwinT, and ResNet-50 actually dropped (DenseNet-201 from 0.5958 ± 0.0062 to 0.5383 ± 0.0553, SwinT from 0.5026 ± 0.1076 to 0.3998 ± 0.0792, and ResNet-50 from 0.7119 ± 0.0529 to 0.4970 ± 0.0563). These results suggest that models trained on suspected PD samples can generalize to confirmed cases better than chance and that multimodal fusion provides meaningful gains for some backbones but not all. Complete performance metrics for this split are shown in **Table 5** and **Figure 6(c)**.

**Table 5.** The performance metrics for all models trained on suspected PD and non-PD data, tested on PD and non-PD data with 500 bootstraps resampling. All values are mean ± standard deviation.

| Model | Data Type | Accuracy | Sensitivity | Specificity | F1 |
| --- | --- | --- | --- | --- | --- |
| ViT | Multimodal | $0.7663 \pm 0.0500$ | $0.5961 \pm 0.0859$ | $0.9694 \pm 0.0319$ | $0.7619 \pm 0.0535$ |
| SwinT | Multimodal | $0.5023 \pm 0.0477$ | $0.0934 \pm 0.0960$ | $0.9902 \pm 0.0182$ | $0.3998 \pm 0.0792$ |
| DenseNet 201 | Multimodal | $0.5886 \pm 0.0383$ | $0.2438 \pm 0.0703$ | $1.0000 \pm 0.0000$ | $0.5383 \pm 0.0553$ |
| ResNet-50 | Multimodal | $0.5610 \pm 0.0365$ | $0.1931 \pm 0.0671$ | $1.0000 \pm 0.0000$ | $0.4970 \pm 0.0563$ |
| MobileNet V2 | Multimodal | $0.6752 \pm 0.0444$ | $0.4778 \pm 0.1149$ | $0.9108 \pm 0.0985$ | $0.6611 \pm 0.0549$ |
| ViT | Images | $0.5415 \pm 0.1018$ | $0.3599 \pm 0.4091$ | $0.7582 \pm 0.4094$ | $0.4263 \pm 0.1504$ |
| SwinT | Images | $0.5686 \pm 0.0702$ | $0.3816 \pm 0.3403$ | $0.7919 \pm 0.2737$ | $0.5026 \pm 0.1076$ |
| DenseNet 201 | Images | $0.6199 \pm 0.0049$ | $0.3544 \pm 0.0089$ | $0.9369 \pm 0.0000$ | $0.5958 \pm 0.0062$ |
| ResNet-50 | Images | $0.7236 \pm 0.0455$ | $0.4921 \pm 0.0836$ | $1.0000 \pm 0.0000$ | $0.7119 \pm 0.0529$ |
| MobileNet V2 | Images | $0.6330 \pm 0.0159$ | $0.3510 \pm 0.0292$ | $0.9695 \pm 0.0000$ | $0.6059 \pm 0.0216$ |
| FFNN | Engineered features | $0.4859 \pm 0.0201$ | $0.0551 \pm 0.0369$ | $1.0000 \pm 0.0000$ | $0.1022 \pm 0.0654$ |

### Feature Importance Analysis of Multimodal Models

We selected the best-performing models from each of the train-test splits based on the highest F1 scores achieved. For the first split, which used only PD and non-PD data for training and testing, the image-based DenseNet-201 model achieved the best performance. For the second and third splits, where suspected PD cases were involved, the best-performing models were the multimodal ResNet-50 and multimodal ViT, respectively. We conducted GradSHAP-based feature importance analysis on these two multimodal models to better understand the relative contributions of image and engineered features. We did not perform feature importance analysis on the image-only DenseNet-201 model, as such an analysis would provide limited additional insight: with no tabular features present, the dominance of image inputs is expected and not subject to comparison. Our goal was to explore how much influence tabular features or image features have in a multimodal setting and whether models rely solely on image inputs or leverage both data types.

**Figure 7** depicts the feature importance analysis for the multimodal ResNet-50 model trained on suspected PD vs non-PD tested on PD vs non-PD. As we observed non-deterministic performance from this model, we selected the best-performing **Figure 7(a)**, median-performing **Figure 7(b)** and the worst-performing **Figure 7(c)** samples from the 20 separate trainings of the model. In all three cases, the model relied predominantly on image features for prediction, with the spiral, sine, and straight-line images showing the highest SHAP values. Among the engineered features, only the time taken to trace the spiral and sine waves as well as the straight-line showed attribution, while the rest of the tabular features had negligible impact. The slight differences in SHAP distributions across the three runs correspond to the relatively small variations in performance, suggesting consistent model behavior.

**Figure 7.**
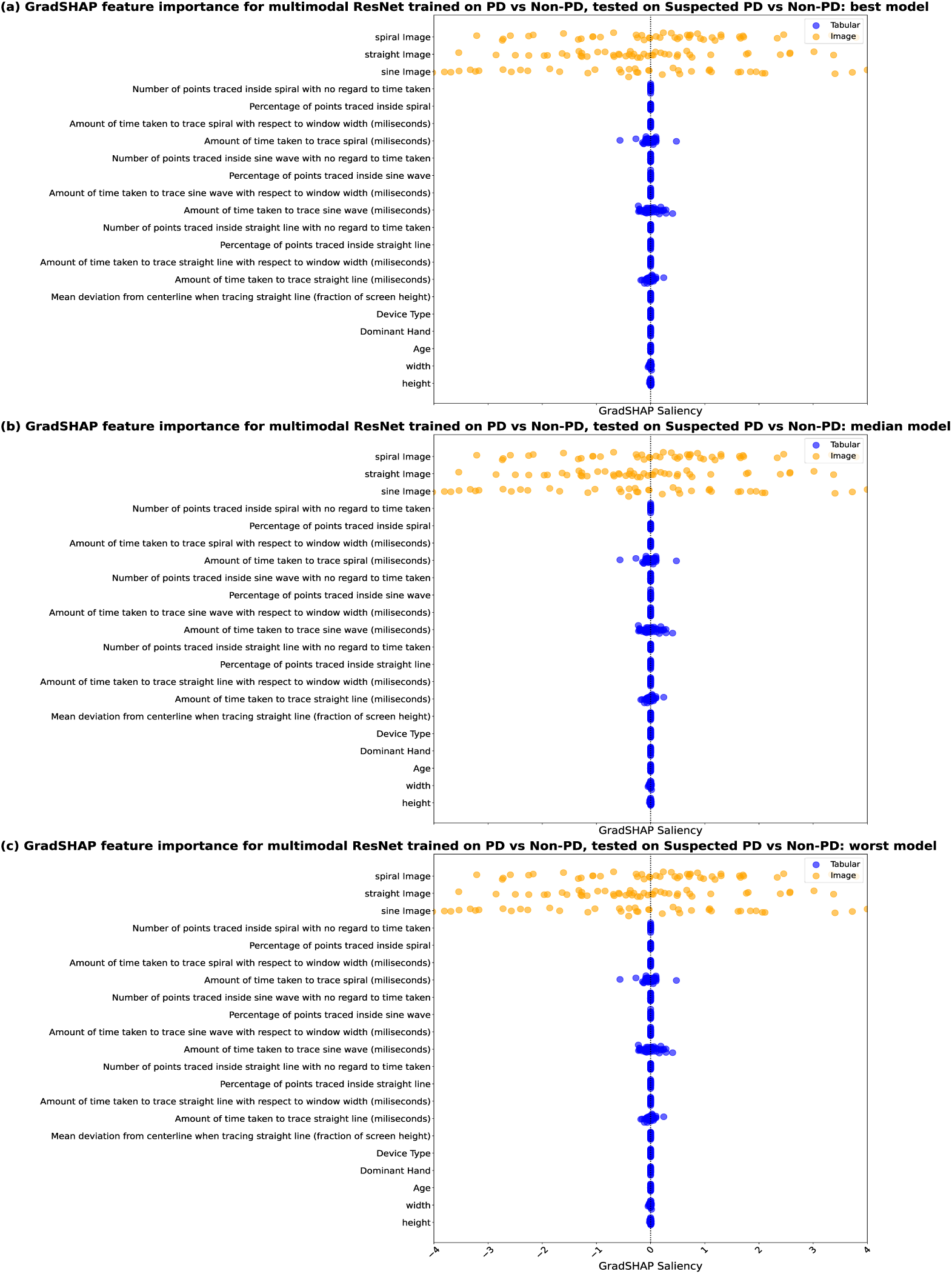
Interpretability analysis of the multimodal ResNet-50 Models for the suspected PD and non-PD cases (trained on PD and non-PD). (a) GradShap attributions showing the feature importance of the best-performing model (b) GradShap attributions showing the feature importance of median model. (c) GradShap attributions showing the feature importance of worst performing. (X-axis range set to (-4,4) for all figures for better comparison and outlier GradShap values removed from figure).

**Figure 8** depicts the feature importance analysis for the multimodal ViT model trained on suspected PD vs. non-PD and tested on PD vs. non-PD. This model showed deterministic performance for this split. So, only one model was selected. The figure shows that the model relied heavily on the images for prediction. However, in this case, the engineered features, especially the amount of time taken to trace the lines, become significantly important with high SHAP values. Also, the height and width of the screen used by the participant also become much important, although to a small extent. This change in attribution of features is important, especially the reliance on the tabular data, which may stem from the training set consisting of suspected PD cases which are not confirmed and can be much more nuanced.

**Figure 8.**
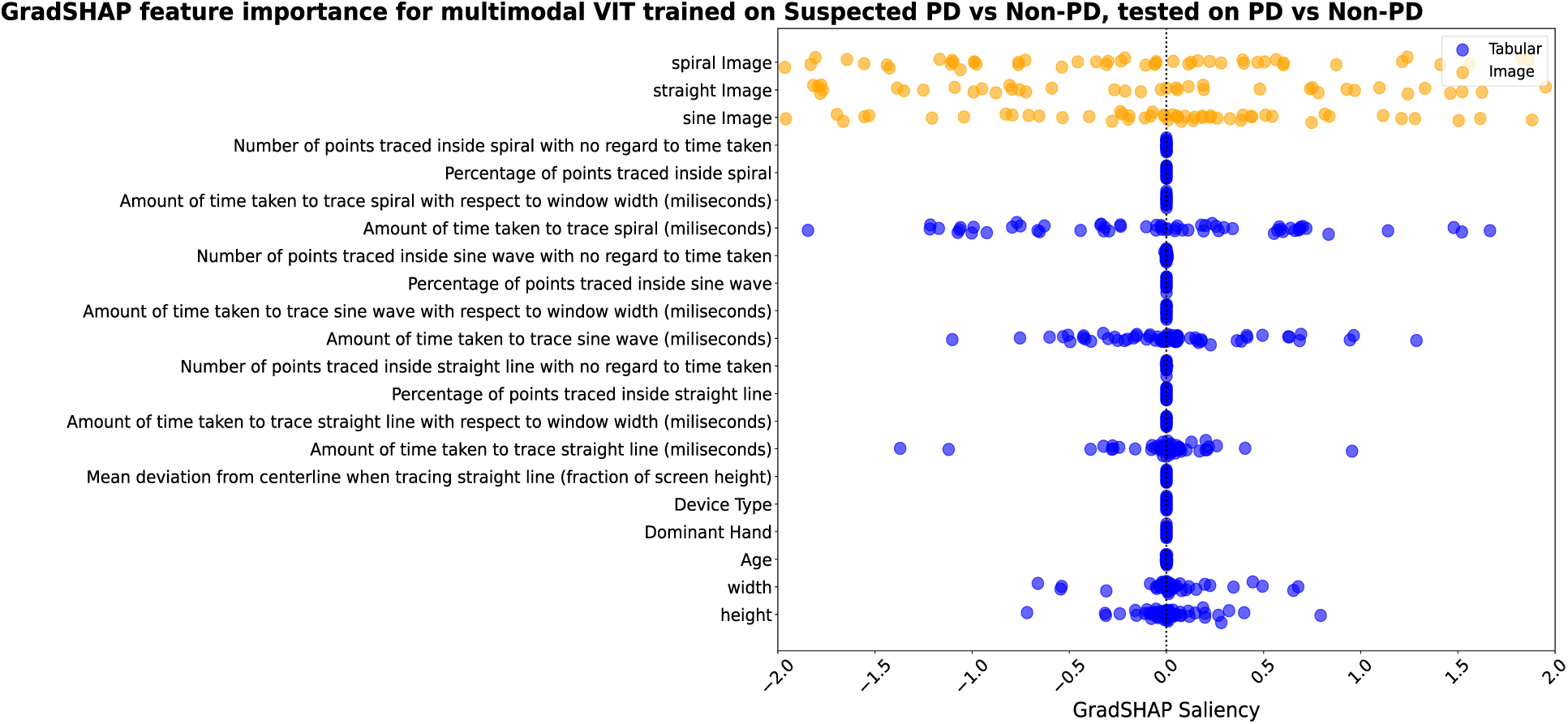
Interpretability analysis of the mulitmodal ViT Model for the PD and non-PD cases (trained on suspected PD and non-PD). (X-axis range set to (-2,2) for the figure for better comparison and outlier GradShap values removed from figure).

## Discussion

### Principal Results

This study demonstrates that multimodal deep learning approaches using mouse trace data can contribute predictive power toward remote PD assessments. We employed three distinct train-test splits in our study. The first split used only confirmed PD and non-PD participants, excluding those with suspected PD. This allowed us to directly assess the diagnostic utility of mouse trace data for differentiating confirmed cases. The second split trained models on confirmed PD and non-PD participants but tested them on suspected PD and non-PD individuals. This served to evaluate whether models trained on verified PD cases could generalize to earlier, potentially subtler motor patterns observed in individuals who self-identify as potentially having PD. The third split reversed this logic: models were trained on suspected PD and non-PD data and tested on confirmed PD cases. This assessed whether self-reported motor symptoms and behaviors in suspected PD could act as a proxy for identifying individuals with actual PD diagnoses.

We explored three different model types and combinations of engineered features and images for multimodal PD detection. For the first and third splits, model performance improved when moving from engineered features to image-based data. But for the second split, we observed improved performance from the tabular model than the image-based models. However, for all the splits, the transition from image-only to multimodal models did not always lead to further improvement. In particular, while some multimodal architectures benefited from incorporating engineered features, others showed limited or inconsistent gains, indicating that only certain architectures effectively leverage multimodal fusion. One likely reason is that most backbone models used in our study were originally developed for image-based tasks and may not optimally integrate tabular features without architectural modifications

Several image-based and multimodal models exhibited non-deterministic behavior during training, likely due to random initialization, dropout, and GPU-level variability (e.g., parallelism, floating-point operation order, and parallel data loading). To ensure model reliability, we conducted 10 independent training runs per fold in the first approach and 20 independent training runs for the other two approaches

Best models were selected based on the highest F1 score achieved across repeated runs in each split. In the first split, the image-only DenseNet-201 achieved the top performance. However, we excluded it from GradSHAP analysis because, as a unimodal model, its feature contributions are limited solely to image data. Since our goal was to compare the relative influence of image versus tabular inputs, we focused GradSHAP analysis on the top-performing multimodal models: ResNet- 50 (split 2) and ViT (split 3). These allowed us to directly assess the role of engineered features in models with both data types.

The analysis showed that image features derived from spiral, sine, and straight-line tracing were the dominant contributors to model predictions in both the multimodal models. Tabular features such as tracing time showed modest attribution in the ResNet-50 (split 2), while showing much higher influence in ViT (split 3). Device-related features (screen height, width) played a minimal to non-existent role. In the ViT model trained on suspected PD cases, the increased influence of engineered features suggests that engineered data can complement visual patterns in using uncertain PD cases to train models to identify real PD cases with more reliability

These findings suggest that multimodal models using models built for image data could be valuable in developing non-invasive, accurate diagnostic tools for PD, facilitating early detection and improved patient management. The interpretability provided by the analyses highlighted that while image data were most important, engineered features played a lesser but important role in predicting the different PD cases. This difference in the importance of image data and engineered features across training and evaluation procedures likely stems from the nature of the data and the specific challenges in each case. For confirmed PD cases, mouse trace images show clearer patterns with respect to non-PD cases, making image features more useful for prediction. In suspected PD cases, the distinctions are more subtle, due to the earlier stage of PD in individuals without an official diagnosis. In this case, the model tends to rely a bit more on engineered features, like tracing time, to capture less obvious differences. This suggests that suspected PD cases require a broader use of data to make accurate predictions. The study also highlights the importance of repeated evaluation, model interpretability, and careful handling of data shifts in developing robust screening tools for PD.

### Comparison to previous works

Our study enhances previous research by introducing an approach to PD detection through the use of multimodal deep learning models, relying exclusively on mouse trace data and images produced from a brief online test. Unlike prior methods that have explored hand and finger movements, keyboard typing patterns, keystroke dynamics, speech analysis, handwriting, drawing tests, and sensor data from accelerometers, gyroscopes, and smartphone interactions, our study focuses specifically on the remote collection of mouse tracing data, demonstrating the potential of mouse trace data alone to provide significant predictive power in predicting PD despite differences in mouse types and devices across participants.

Previous studies by Gil-Martin et al.^34^ and Pereira et al.^35^ focused on hand movement dynamics from spiral, meander, and other drawing shapes for PD analysis. However, their data collection was not remote, and they did not consider handedness, unlike our study. Their best models achieved accuracies of 97.7% and 83.77%, respectively. Goel et al.^36^ used pen-and-paper methods to collect spiral pattern data but also lacked remote testing and consideration of handedness, achieving an accuracy of 84.73%. Memedi et al.^37^ used a remote data collection method involving a touchscreen tablet and web interface, but their study spanned three years and involved only 65 participants, resulting in an accuracy of around 84.73%. Wang et al.^38^ achieved 96.2% accuracy using data collected during the drawing of Π and ΠΛ shapes. However, their data collection method involved an iPad and stylus and was not conducted remotely. Classification model suggested by Gajos et al.^39^ detected ataxia or parkinsonism with high sensitivity (0.91) and specificity (0.90) but had only 46 patients with PD in their data set with 22906 controls.

While our study differs significantly from these prior works in terms of data collection methods, the duration of data collection, remote accessibility, and the inclusion of handedness, these studies serve as important foundational works.

Our pilot study^40^, which explored the feasibility of an earlier version of our web application, achieved an accuracy of 74.29% and an F1 score of 73.11%, but used keyboard test data along with the mouse trace data. Our current model shows marked improvements in performance, reflecting the advancements and refinements made in our approach.

### Limitations & Future work

This study has several limitations that should be considered in future work. First, the sample size of 261 split between three diagnostic categories may not be sufficient to generalize the findings across a diverse population. Moreover, while we performed hyperparameter tuning for each model, additional optimization, especially regarding model architecture and data fusion strategies, could further improve performance and generalization. Some models, particularly those involving image-based or multimodal inputs, exhibited non-deterministic behavior across repeated runs on the same data splits. These models produced variable results across repeated training runs on identical data splits, likely due to the stochastic nature of deep learning and their sensitivity to complex, high-dimensional input spaces. This reinforces the importance of repeated experimentation and performance averaging in evaluating model robustness. We used 500 ms differences between each recorded point, while participants traced the lines. This resulted in one or two cases each for the straight-line, sine-wave and spiral-wave to record a single point of data. We kept those data in this study, but we also plan to record the points more frequently (e.g. 50/100 ms) to overcome this.

Additionally, our focus on mouse tracing data collected through a website does not fully capture all aspects of PD symptoms. Importantly, the study did not count medication usage, specifically accounting for the on phase versus the off phase, which can significantly influence the presence and severity of symptoms like tremors^41,42,43^. Stress, which is known to exacerbate tremors, was also not accounted for, potentially affecting the results^44^. The type of mouse used by participants, such as whether they used an ergonomic mouse or a computer trackpad, could have influenced the prominence of tremor symptoms, introducing variability in the data. Furthermore, the impact of device type and handedness on the results remains unclear, as PD often affects one side of the body more than the other, and it is not certain that participants’ dominant hands were the ones most affected by the disease. While the transformer-based models demonstrated a strong performance, their computational complexity and resource requirements may limit their practical application in real-world settings. Future research should focus on optimizing these models for use on standard hardware without compromising performance and should incorporate additional data modalities, such as voice recordings and gait analysis, to provide a more comprehensive diagnostic approach.

## Data Availability

An anonymized version of the data used in this study may be released upon completion of the ongoing data collection process.

## Contributors

Conceptualization: PW, RSZ; Data collection: RSZ, ZNT, LS, SP; Web development: RSZ, SP, LS; Writing - primary writing: RSZ; Writing - editing: PW; Funding acquisition: PW; Data analysis: RSZ; Visualization: RSZ; Ideation: RSZ, LS, ZNT, SP, PW; Supervision: PW. All authors had full access to the data used in the study and had final responsibility for the decision to submit for publication.

## Code Sharing

The code for this study [and training/validation datasets] are not publicly available but may be made available to researchers on reasonable request to the first author.

## Acknowledgements

The authors are grateful to the participants who participated in this study. This research was, in part, funded by the National Institutes of Health (NIH) Agreement NO. 1OT2OD032581-01. The views and conclusions contained in this document are those of the authors and should not be interpreted as representing the official policies, either expressed or implied, of the NIH. Only the language and grammar of this manuscript was revised using AI tools such as ChatGPT, though the output of the AI tools was further edited by the authors.

## References

1. Burke RE. Evaluation of the Braak staging scheme for Parkinson’s disease: Introduction to a panel presentation. Movement Disorders. 2010;25(S1):S76–7.

2. Olanow CW, Stern MB, Sethi K. The scientific and clinical basis for the treatment of Parkinson disease (2009). Neurology [Internet]. 2009 May 26 [cited 2024 Jul 17];72(21_supplement_4). Available from: https://www.neurology.org/doi/10.1212/WNL.0b013e3181a1d44c

3. What is Parkinson’s? | Parkinson’s Foundation [Internet]. [cited 2024 Jul 17]. Available from: https://www.parkinson.org/understanding-parkinsons/what-is-parkinsons

4. Statistics | Parkinson’s Foundation [Internet]. [cited 2024 Jul 17]. Available from: https://www.parkinson.org/understanding-parkinsons/statistics

5. Pangman VC, Sloan J, Guse L. An examination of psychometric properties of the mini-mental state examination and the standardized mini-mental state examination: implications for clinical practice. Appl Nurs Res. 2000 Nov;13(4):209–13.

6. Hamzehei S, Akbarzadeh O, Attar H, Rezaee K, Fasihihour N, Khosravi MR. Predicting the total Unified Parkinson’s Disease Rating Scale (UPDRS) based on ML techniques and cloud-based update. Journal of Cloud Computing. 2023 Jan 21;12(1):12.

7. Myrberg K, Hydén LC, Samuelsson C. The mini-mental state examination (MMSE) from a language perspective: an analysis of test interaction. Clinical Linguistics & Phonetics. 2020 Jul 2;34(7):652–70.

8. Predicting Early Stage Drug Induced Parkinsonism using Unsupervised and Supervised Machine Learning | IEEE Conference Publication | IEEE Xplore [Internet]. [cited 2024 Mar 13]. Available from: https://ieeexplore.ieee.org/document/9175343

9. How Parkinson’s disease is diagnosed. American Parkinson Disease Association. URL: https://www.apdaparkinson.org/what-is-parkinsons/diagnosing/ [accessed 2024-09-21]

10. How is Parkinson’s diagnosed? Parkinson’s Europe. URL: https://www.parkinsonseurope.org/about-parkinsons/diagnosis/how-is-parkinsons-diagnosed/ [accessed 2024-09-21]

11. Dopaminergic neuron-specific oxidative stress caused by dopamine itself - PubMed [Internet]. [cited 2024 Jul 18]. Available from: https://pubmed.ncbi.nlm.nih.gov/18596830/

12. Rovini E, Maremmani C, Cavallo F. How Wearable Sensors Can Support Parkinson’s Disease Diagnosis and Treatment: A Systematic Review. Front Neurosci [Internet]. 2017 Oct 6 [cited 2024 Jul 19];11. Available from: https://www.frontiersin.org/journals/neuroscience/articles/10.3389/fnins.2017.00555/full

13. Arroyo-Gallego T, Ledesma-Carbayo MJ, Butterworth I, Matarazzo M, Montero-Escribano P, Puertas-Martín V, et al. Detecting Motor Impairment in Early Parkinson’s Disease via Natural Typing Interaction With Keyboards: Validation of the neuroQWERTY Approach in an Uncontrolled At-Home Setting. J Med Internet Res. 2018 Mar 26;20(3):e89.

14. Demir B, Ulukaya S, Erdem O. Detection of Parkinson’s disease with keystroke data. Comput Methods Biomech Biomed Engin. 2023 Oct;26(13):1653–67.

15. Tsanas A, Little MA, McSharry PE, Ramig LO. Accurate telemonitoring of Parkinson’s disease progression by noninvasive speech tests. IEEE Trans Biomed Eng. 2010 Apr;57(4):884–93.

16. Del Din S, Elshehabi M, Galna B, Hobert MA, Warmerdam E, Suenkel U, et al. Gait analysis with wearables predicts conversion to parkinson disease. Ann Neurol. 2019 Sep;86(3):357–67.

17. Klucken J, Barth J, Kugler P, Schlachetzki J, Henze T, Marxreiter F, et al. Unbiased and mobile gait analysis detects motor impairment in Parkinson’s disease. PLoS One. 2013;8(2):e56956.

18. Cancela J, Pastorino M, Tzallas AT, Tsipouras MG, Rigas G, Arredondo MT, et al. Wearability Assessment of a Wearable System for Parkinson’s Disease Remote Monitoring Based on a Body Area Network of Sensors. Sensors (Basel). 2014 Sep 16;14(9):17235–55.

19. Patel S, Lorincz K, Hughes R, Huggins N, Growdon J, Standaert D, et al. Monitoring motor fluctuations in patients with Parkinson’s disease using wearable sensors. IEEE Trans Inf Technol Biomed. 2009 Nov;13(6):864–73.

20. Giancardo L, Sánchez-Ferro A, Arroyo-Gallego T, Butterworth I, Mendoza CS, Montero P, et al. Computer keyboard interaction as an indicator of early Parkinson’s disease. Sci Rep. 2016 Oct 5;6(1):34468.

21. Wylie SA, Van Den Wildenberg WP, Ridderinkhof KR, Bashore TR, Powell VD, Manning CA, Wooten GF. The effect of Parkinson’s disease on interference control during action selection. Neuropsychologia. 2009 Jan 1;47(1):145–57.

22. Goñi M, Eickhoff SB, Far MS, Patil KR, Dukart J. Smartphone-based digital biomarkers for Parkinson’s disease in a remotely-administered setting. IEEE access. 2022 Mar 3;10:28361–84.

23. Skaramagkas V, Andrikopoulos G, Kefalopoulou Z, Polychronopoulos P. A study on the essential and Parkinson’s arm tremor classification. Signals. 2021 Apr 19;2(2):201–24.

24. Schneider SA, Drude L, Kasten M, Klein C, Hagenah J. A study of subtle motor signs in early Parkinson’s disease. Movement disorders. 2012 Oct;27(12):1563–6.

25. Prince J, Andreotti F, De Vos M. Multi-Source Ensemble Learning for the Remote Prediction of Parkinson’s Disease in the Presence of Source-Wise Missing Data. IEEE Trans Biomed Eng. 2019 May;66(5):1402–11.

26. Vasquez-Correa JC, Arias-Vergara T, Orozco-Arroyave JR, Eskofier B, Klucken J, Noth E. Multimodal Assessment of Parkinson’s Disease: A Deep Learning Approach. IEEE J Biomed Health Inform. 2019 Jul;23(4):1618–30.

27. Kandori A, Yokoe M, Sakoda S, Abe K, Miyashita T, Oe H, et al. Quantitative magnetic detection of finger movements in patients with Parkinson’s disease. Neuroscience Research. 2004 Jun 1;49(2):253–60.

28. Bhattacharjee M, Bandyopadhyay D. Flexible Paper Touchpad for Parkinson’s Hand Tremor Detection. Sensors and Actuators A: Physical. 2019 Aug 1;294:164–72.

29. Lakshminarayana R, Wang D, Burn D, Chaudhuri KR, Galtrey C, Guzman NV, et al. Using a smartphone-based self-management platform to support medication adherence and clinical consultation in Parkinson’s disease. NPJ Parkinsons Dis. 2017 Nov 13;3:2.

30. Lipsmeier F, Taylor KI, Kilchenmann T, Wolf D, Scotland A, Schjodt-Eriksen J, et al. Evaluation of smartphone-based testing to generate exploratory outcome measures in a phase 1 Parkinson’s disease clinical trial. Mov Disord. 2018 Aug;33(8):1287–97.

31. Wan S, Liang Y, Zhang Y, Guizani M. Deep Multi-Layer Perceptron Classifier for Behavior Analysis to Estimate Parkinson’s Disease Severity Using Smartphones. IEEE Access. 2018 Jul 6;PP:1–1.

32. Li W, Zhu W, Dorsey ER, Luo J. Predicting Parkinson’s Disease with Multimodal Irregularly Collected Longitudinal Smartphone Data [Internet]. arXiv; 2020 [cited 2024 Jul 19]. Available from: http://arxiv.org/abs/2009.11999

33. Lundberg S. A unified approach to interpreting model predictions. arXiv preprint arXiv:1705.07874. 2017.

34. Gil-Martín M, Montero JM, San-Segundo R. Parkinson’s Disease Detection from Drawing Movements Using Convolutional Neural Networks. Electronics. 2019 Aug 17;8(8):907.

35. Pereira CR, Weber SAT, Hook C, Rosa GH, Papa JP. Deep Learning-Aided Parkinson’s Disease Diagnosis from Handwritten Dynamics. In: 2016 29th SIBGRAPI Conference on Graphics, Patterns and Images (SIBGRAPI) [Internet]. Sao Paulo, Brazil: IEEE; 2016 [cited 2024 Aug 28]. p. 340–6. Available from: http://ieeexplore.ieee.org/document/7813053/

36. Goel N, Khanna A, Gupta D, Gupta N. Detection of Parkinson’s Disease Using Machine Learning Techniques for Voice and Handwriting Features. In: Khanna A, Gupta D, Bhattacharyya S, Snasel V, Platos J, Hassanien AE, editors. International Conference on Innovative Computing and Communications. Singapore: Springer; 2020. p. 631–43.

37. Memedi M, Sadikov A, Groznik V, Žabkar J, Možina M, Bergquist F, et al. Automatic Spiral Analysis for Objective Assessment of Motor Symptoms in Parkinson’s Disease. Sensors. 2015 Sep;15(9):23727–44.

38. Wang X, Huang J, Chatzakou M, Medijainen K, Toomela A, Nõmm S, Ruzhansky M. LSTM-CNN: An efficient diagnostic network for Parkinson’s disease utilizing dynamic handwriting analysis. Computer Methods and Programs in Biomedicine. 2024 Apr 1;247:108066.

39. Gajos KZ, Reinecke K, Donovan M, Stephen CD, Hung AY, Schmahmann JD, Gupta AS. Computer mouse use captures ataxia and parkinsonism, enabling accurate measurement and detection. Movement Disorders. 2020 Feb;35(2):354–8.

40. Parab S, Boster JR, Washington P. Parkinson’s Disease Recognition using a Gamified Website: Machine Learning Feasibility Study [Internet]. medRxiv; 2023 [cited 2023 Dec 22]. p. 2023.08.22.23294440. Available from: https://www.medrxiv.org/content/10.1101/2023.08.22.23294440v1

41. Sieberts SK, Schaff J, Duda M, Pataki BÁ, Sun M, Snyder P, et al. Crowdsourcing digital health measures to predict Parkinson’s disease severity: the Parkinson’s Disease Digital Biomarker DREAM Challenge. npj Digit Med. 2021 Mar 19;4(1):1–12.

42. Severson KA, Chahine LM, Smolensky LA, Dhuliawala M, Frasier M, Ng K, et al. Discovery of Parkinson’s disease states and disease progression modelling: a longitudinal data study using machine learning. The Lancet Digital Health. 2021 Sep 1;3(9):e555–64.

43. Roussos G, Herrero TR, Hill DL, Dowling AV, LTM Müller M, Evers LJ, Burton J, Derungs A, Fisher K, Kilambi KP, Mehrotra N. Identifying and characterising sources of variability in digital outcome measures in Parkinson’s disease. NPJ Digital Medicine. 2022 Jul 15;5(1):93.

44. Rochester L, Hetherington V, Jones D, Nieuwboer A, Willems AM, Kwakkel G, Van Wegen E. Attending to the task: interference effects of functional tasks on walking in Parkinson’s disease and the roles of cognition, depression, fatigue, and balance. Archives of physical medicine and rehabilitation. 2004 Oct 1;85(10):1578–85.

